# The *GBA1* p.E427K (p.E388K) Variant is a Risk Factor for Synucleinopathies: A Meta-Analysis

**DOI:** 10.64898/2026.03.18.26347268

**Authors:** Leah V. Chifamba, Sitki Cem Parlar, Emma N. Somerville, Lang Liu, Eric Yu, Farnaz Asayesh, Jamil Ahmad, Meron Teferra, Jennifer A. Ruskey, Cheryl Waters, Oury Monchi, Yves Dauvilliers, Nicolas Dupré, Alla Timofeeva, Anton Emelyanov, Sofya Pchelina, Irina Miliukhina, Lior Greenbaum, Sharon Hassin-Baer, Orly Goldstein, Mandy Radefeldt, Peter Bauer, Christian Beetz, Allison A. Dilliott, James C. Beck, Konstantin Senkevich, ROPAD Study Group, The Parkinson’s Foundation PD GENEration Study, Christine Klein, Roy N. Alcalay, Ziv Gan-Or

## Abstract

**Background:** Variants in *GBA1* are important genetic risk factors for synucleinopathies, including Parkinson’s disease (PD). While several *GBA1* variants are established risk or severity modifiers, the role of the p.E427K variant remains unclear.

Objective

To determine whether the *GBA1* p.E427K variant is associated with risk of synucleinopathies.

**Methods:** We performed a meta-analysis of case-control studies reporting the frequency of *GBA1* p.E427K (p.E388K) in PD and related synucleinopathies. Data were obtained from published studies, open-access resources, and large cohorts, including in-house datasets. Odds ratios (ORs) were calculated for each cohort and pooled using a random-effects model.

**Results:** Across 67,484 patients and 124,079 controls, *GBA1* p.E427K was associated with increased disease risk (pooled OR = 1.87, 95% CI 1.28–2.72, P = 0.001). Enzymatic data showed reduced glucocerebrosidase activity in carriers.

**Conclusions:** The *GBA1* p.E427K variant is a risk factor for synucleinopathies and should be considered in genetic studies and clinical trials.

## Introduction

*GBA1* variants are important genetic risk factors for synucleinopathies and are found in approximately 5-33% of patients with Parkinson’s disease (PD), dementia with Lewy bodies (DLB), or the prodromal condition rapid eye movement (REM) sleep behavior disorder (RBD) (1–3). Given their high prevalence, clinical trials are currently recruiting individuals with *GBA1* variants, with additional trials planned in the future (4).

The classification of *GBA1* variants is critical for such trials, as different types of *GBA1* variants may be associated with distinct rates of disease progression (5). Based on their phenotypic effects in homozygous state in Gaucher disease (GD), *GBA1* variants can be classified as *mild* if they cause GD type 1, or *severe* if they cause GD types 2 or 3 (6). An additional category includes *GBA1* variants that do not cause GD but are associated with an increased risk of PD, and these are classified as *risk variants* (6). Carriers of severe *GBA1* variants have an earlier age at onset and experience more rapid progression of both motor and non-motor symptoms (7). Therefore, to ensure balanced progression rates in clinical trial arms, an equal distribution of severe, mild and risk *GBA1* variants across treatment and placebo groups is required.

Currently, three *GBA1* variants are reliably classified as risk factors: p.E365K (p.E326K), p.T408M (p.T369M) (8), and the African-specific intronic splicing variant rs3115534-G (9). Another variant, p.E427K (p.E388K), has been proposed as a risk factor for PD (1). However, this association has not been confirmed by robust genetic analyses. Consequently, carriers of p.E427K were not included in the first two phase 2 clinical trials targeting PD patients with *GBA1* variants (https://clinicaltrials.gov/study/NCT05819359) (10).

In the present study, we evaluated the role of *GBA1* p.E427K in synucleinopathies using data from 67,484 patients and 124,079 controls across multiple cohorts, followed by a meta-analysis of effects across cohorts.

## Subjects and Methods

### Data sources and cohorts

We identified published case-control studies reporting genotyping data for *GBA1* p.E427K (also termed p.E388K, rs149171124) variant in PD cohorts using the GBA1-PD browser (6) (accessed October 2025). Studies were eligible for inclusion if they reported the number of p.E427K carriers among PD cases and controls. Studies without available carrier counts or with overlapping cohorts were excluded. Using these criteria, seven studies were included (12,442 PD cases and 6,001 controls) (Table 1).

**Table 1:** Distribution of *GBA1* p.E388K carriers across cohorts included in the meta-analysis.

| Cohort | Synucleinopathy | Carriers | Carriers | Total | Total |
| --- | --- | --- | --- | --- | --- |
|  |  | Cases | Controls | Cases | Controls |
| Parkinson Institute Biobank (28) | PD | 5 | 1 | 2,766 | 1,111 |
| European-American (29) | PD | 1 | 0 | 478 | 337 |
| Dutch (30) | PD | 3 | 0 | 3,402 | 655 |
| Not specified (31) | PD | 1 | 0 | 185 | 257 |
| European (32) | PD | 1 | 0 | 1,130 | 391 |
| Colombian and Peruvian (33) | PD | 0 | 1 | 602 | 319 |
| Chinese (34) | PD | 1 | 0 | 3,879 | 2,931 |
| AMP-PD | DLB | 2 | 0 | 2,605 | 1,894 |
| AMP-PD | PD | 1 | 2 | 1,820 | 2,824 |
| Centogene (ROPAD) | PD | 30 | 21 | 20,102 | 35,134 |
| GP2 | PD | 6 | 5 | 17,998 | 15,760 |
| McGill | PD | 10 | 3 | 7,111 | 5,493<sup>1</sup> |
| McGill | RBD | 2 | 3 | 2,800 | 5,493 |
| UKBB | PD | 2 | 19 | 2,606 | 56,973 |
| Total |  | 65 | 52 | 67,484 | 124,079 |

PD, Parkinson's disease; DLB, dementia with Lewy bodies; RBD, rapid eye movement sleep behavior disorder; UKBB, UK Biobank; AMP-PD, Accelerating Medicines Partnership–Parkinson's Disease; GP2, Global Parkinson's Genetics Program; ROPAD, Rostock International Parkinson's Disease Study; N, number of individuals
<sup>1</sup> McGill controls (N=5,493) are shared between the McGill PD and McGill RBD comparisons; controls were counted once in the overall totals

Additional data were obtained from several cohorts, including: (i) In-house McGill University cohorts comprising 7,111 PD patients, 2,800 RBD patients and 5,493 controls. The cohorts were recruited in Quebec, Canada and Montpellier, France (11), Columbia University, New York (12), Sheba Medical Center, Israel (13) and Pavlov First State Medical University and the Institute of Human Brain, Russia (14). (ii) the Centogene - Rostock International Parkinson’s Disease Study (ROPAD) (15), comprising 20,102 patients with PD and 35,134 controls (16); (iii) the PD GENEration (17) with 25,932 PD patients and no available control group, therefore, this cohort was not included in the meta-analysis; (iv) Accelerated Medicines Partnership - Parkinson’s Disease (AMP-PD) (https://amp-pd.org/), including 1,820 PD, 2,605 DLB cases and 4,718 controls; (v) the UK Biobank (UKBB) (https://www.ukbiobank.ac.uk/), with 2,606 PD patients and 56,973 controls; and (vi) data from Global Parkinson’s Genetics Program (GP2) (18), consisting of 17,998 cases and 15,760 controls, obtained from the publicly available browser, (https://gp2.broadinstitute.org/) (18).

### Genetic sequencing and quality control

Targeted next-generation sequencing of *GBA1* in the McGill University cohorts was performed using molecular inversion probes, as previously described (19). Sequencing was conducted at the McGill Genome Center using the Illumina NovaSeq 6000 SP PE100 platform. Reads were aligned to the hg19 reference genome using the Burrows-Wheeler Aligner, and post-alignment processing and variant calling were performed with Genome Analysis Toolkit (GATK v3.8). Quality control (QC) was carried out using PLINK v1.9 (20), excluding variants with missingness of >10%, genotype quality (GQ) < 30, or sequencing depth < 30x.

Whole-genome sequencing (WGS) data for the AMP-PD and UKBB cohorts were obtained from Terra platform (https://app.terra.bio/) and UKBB Research Analysis Platform (https://www.ukbiobank.ac.uk/) respectively. AMP-PD data underwent sample and variant-level QC as previously described (21), while UK Biobank WGS data were processed using GATK v3.8, applying a minimum sequencing depth coverage of 30x and GQ ≥20. The hg38 reference genome was used for both AMP-PD and UK Biobank datasets.

For Global Parkinson’s Genetics Program (GP2), variant carrier counts for *GBA1* p.E427K were obtained from the GP2 Genome Browser (18), an open-access resource aggregating harmonized summary-level genotyping data from multiple contributing cohorts, using the most recent version (September 2025). Only summary-level data were accessed, and no individual-level sequencing data or additional QC procedures were performed by the authors.

*GBA1* p.E427K variant data from the Centogene ROPAD cohort were generated using panel sequencing, whole-exome sequencing (WES), and/or WGS, as previously described (16). Similarly, PD GENEration cohort data were sequenced using WES and WGS (17). In both cohorts, processed, high-quality variant calls were provided by the respective study teams, and no additional sequencing or QC procedures were performed in the present study. All participating institutions received institutional review board approval, and all participants provided written informed consent.

### Variant definition

The *GBA1* p.E427K is referred to using the nomenclature including the 39-amino acid signal peptide of glucocerebrosidase. The alternative nomenclature p.E388K, which excludes the signal peptide, is provided in parentheses where relevant. All cohorts were harmonized to this nomenclature prior to statistical analysis.

### Meta-analysis

All cohorts included in the meta-analysis provided carrier counts for both cases and controls. For each study and cohort, a 2 × 2 contingency table was constructed comparing the number of carriers of *GBA1* p.E427K among patients with synucleinopathies and controls. Odds ratios (ORs) with 95% confidence intervals (CIs) were calculated from these tables. The standard error of the log(OR) was derived using the inverse variance method. To account for variability across studies, pooled effect estimates were obtained using a random-effects model. Between-study heterogeneity was assessed using Cochran’s Q statistic and quantified with the I² index. A constant continuity correction of 0.5 was applied in studies or cohorts where zero counts were present in either the case or control arm, allowing OR calculations. Forest plots were generated in Python using the matplotlib package (22).

## Results

We examined the association between *GBA1* p.E427K and synucleinopathies across all included studies and cohorts using a random-effects meta-analysis (Figure 1A). In the pooled analysis, carriage of *GBA1* p.E427K was associated with an increased risk of synucleinopathies (pooled OR = 1.87, 95% CI = 1.28–2.72, P = 0.001). There was no evidence of between study heterogeneity (Q = 9.20, degrees of freedom [df] = 15, P = 0.87; I² = 0%). We repeated the analysis without the data from GP2 in case there is sample overlap with ROPAD or other studies and received similar results (Figure 1B). Glucocerebrosidase enzymatic activity data were available for 11 heterozygous p.E427K carriers from Centogene and were, on average, 32% lower than in non-carriers (p = 0.0015).

**Figure 1A:**
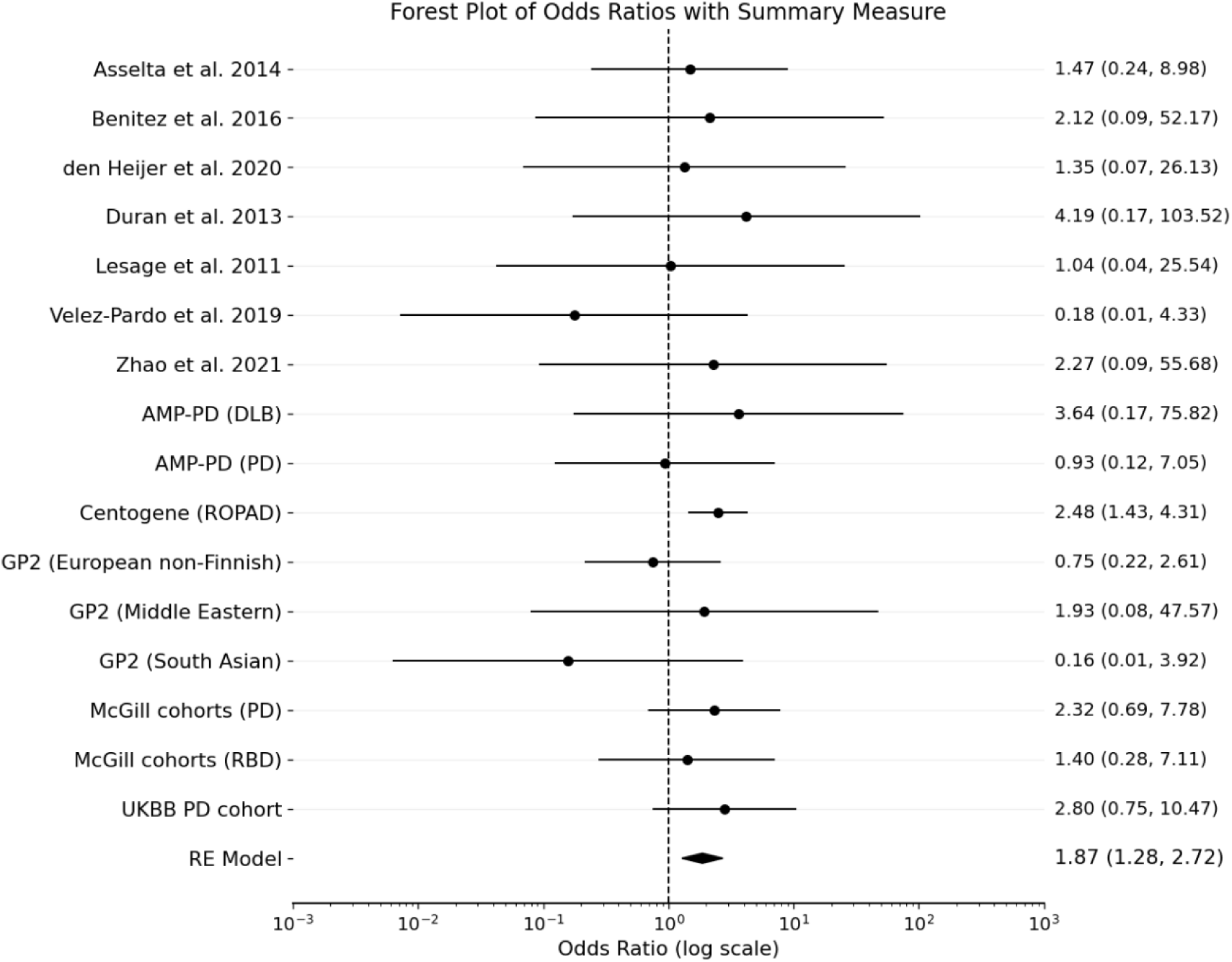
Forest plot of studies with available case control data on the *GBA1* p.E388K variant across cohorts. RE, random effects; PD, Parkinson’s disease; DLB, dementia with Lewy bodies; RBD, rapid eye movement sleep behavior disorder; UKBB, UK Biobank; AMP-PD, Accelerating Medicines Partnership–Parkinson’s Disease; GP2, Global Parkinson’s Genetics Program; ROPAD, Rostock International Parkinson’s Disease Study; N, number of individuals

**Figure 1B:**
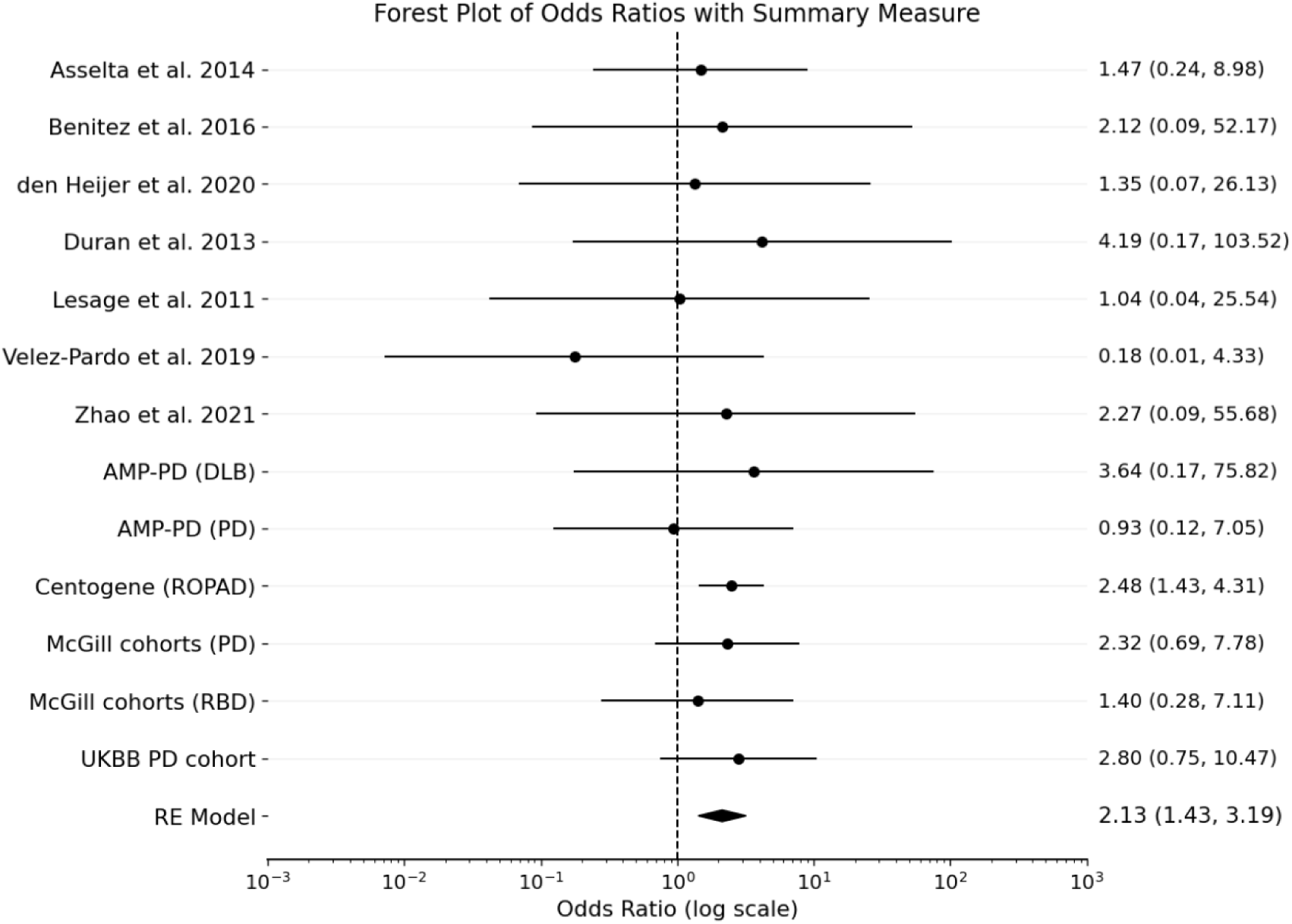
Forest plot of studies with available case control data on the *GBA1* p.E388K variant across cohorts without GP2 cohorts. RE, random effects; PD, Parkinson’s disease; DLB, dementia with Lewy bodies; RBD, rapid eye movement sleep behavior disorder; UKBB, UK Biobank; AMP-PD, Accelerating Medicines Partnership–Parkinson’s Disease; ROPAD, Rostock International Parkinson’s Disease Study; N, number of individuals

In addition to the case-control datasets included in the meta-analysis, we examined the frequency and ethnicity of *GBA1* p.E427K in the PD GENEration cohort, a large independent case-only sequencing dataset. Among 25,932 individuals with PD, 13 heterozygous carriers of *GBA1* p.E427K were identified, including individuals of European, Hispanic/Latino, Black and Ashkenazi Jewish ancestries, suggesting that this variant is relevant across multiple populations. Indeed, in gnomAD (https://gnomad.broadinstitute.org/), this variant is reported with similar frequencies in Europeans and South Asians, with lower frequencies in Finnish Europeans, Admixed Americans, and African/African American.

## Discussion

In the current study, analysing a total of 67,484 patients and 124,079 controls from the literature, open-access resources, and in-house cohorts, we demonstrate that the *GBA1* p.E427K is a risk factor for synucleinopathies, with an effect size comparable to that of the three other established PD risk variants, p.E365K, p.T408M and rs3115534.

Similar to p.E365K, which has an estimated OR for PD risk of 1.8–2.0 (23), and p.T408M, which has an OR of 1.7–1.9 (8,23), the OR for p.E427K in our meta-analysis was ∼1.8. Since p.E427K is less frequent than p.E365K and p.T408M in the general population (allele frequency in Europeans in gnomAD of 0.0002 vs. 0.013 and 0.008, respectively), which explains why previous studies examining this variant were underpowered to detect an association with PD (23). The reduction of GCase activity in the 11 samples available from Centogene is comparable to that reported for carriers of p.E365K and p.T408M (24), further supporting p.E427K as a PD risk variant.

One PD patient in our study was homozygous for the *GBA1* p.E427K variant and, in their early 60s, exhibited no signs or symptoms of GD. This individual had mildly elevated lyso-Gb1 (glucosylsphingosine) levels (9.7 ng/ml) compared with controls (4–7 ng/ml), but substantially lower than levels typically observed in GD patients (>20 ng/ml, often >100 ng/ml). A previous study identified two sisters who were compound heterozygous for *GBA1* p.N409S/p.E427K and reported that both were asymptomatic, although they were very young at the time of evaluation (25). One study of Colombian GD patients reported an individual homozygous for the p.E427K variant. However, this report cites another Colombian study (26), as having identified the same variant in Colombian patients with PD, which is not the case. That study did not report the presence of this variant. Therefore, the purported association between homozygous *GBA1* p.E427K and GD should be interpreted with caution. Based on the currently available data, there is no reliable evidence that this variant causes GD. Moreover, given the variant’s frequency in European populations (∼0.0002), one would expect to observe GD patients homozygous for this variant if it were pathogenic, particularly since much rarer variants, such as p.L483P and others, have been reported repeatedly in patients with GD (27).

A key limitation of this study is the small number of variant carriers within individual cohorts, often ranging from 0–5 per study arm. As a result, single-cohort effect estimates may be unstable and highly sensitive to minor differences in QC or ancestry classification. Although a random-effects meta-analysis model was applied to account for between-study variability, effect sizes from individual cohorts should be interpreted with caution. The pooled meta-analytic estimate therefore represents the most reliable and appropriate reference for inference, rather than any single-study result.

In conclusion, our study demonstrates that *GBA1* p.E427K is a risk factor for synucleinopathies. Accordingly, carriers of this variant can be included in clinical studies and therapeutic trials in a manner similar to carriers of the established *GBA1* risk variants p.E365K and p.T408M.

## Acknowledgments

We acknowledge the participants from the contributing cohorts whose involvement made this study possible. This work was supported in part by the Canada First Research Excellence Fund through McGill University’s Healthy Brains, Healthy Lives initiative, with additional computational support from Calcul Québec and Compute Canada. Z.G.-O. receives support from the Fonds de Recherche du Québec–Santé through the Chercheurs-Boursiers Award, in partnership with Parkinson Quebec, and holds a William Dawson Scholar appointment. Participant recruitment for this study was facilitated in part by the Quebec Parkinson’s Network (http://rpq-qpn.ca/en/). Access to UK Biobank data was enabled by the NeuroHub infrastructure under Application Number 45551 and was undertaken thanks in part to funding from the Canada First Research Excellence Fund. Which was awarded through the Healthy Brains, Healthy Lives initiative at McGill University, and was enabled in part by support provided by Calcul Québec and the Digital Research Alliance of Canada. Furthermore, data used in this article was also obtained in January 2023 from the Accelerating Medicines Partnership® (AMP®) Parkinson’s Disease (AMP PD) Knowledge Platform, release 2.5. More recent releases have since become available. For the latest updates on AMP-PD, please visit https://www.amp-pd.org. The AMP® PD program is a public-private partnership managed by the Foundation for the National Institutes of Health and funded by the National Institute of Neurological Disorders and Stroke (NINDS) in partnership with the Aligning Science Across Parkinson’s (ASAP) initiative; Celgene Corporation, a subsidiary of Bristol-Myers Squibb Company; GlaxoSmithKline plc (GSK); The Michael J. Fox Foundation for Parkinson’s Research; Pfizer Inc.; AbbVie Inc.; Sanofi US Services Inc.; and Verily Life Sciences.

Additional Data used in the preparation of this article were obtained from the Parkinson’s Foundation PD GENEration study. PD GENEration is a flagship initiative of the Parkinson’s Foundation in partnership with Aligning Science Across Parkinson’s (ASAP) and the Global Parkinson’s Genetics Program (GP2). Additional financial and in-kind support comes from the Parkinson’s community – industry partners, nonprofit organizations, and individuals whose lives have been touched by Parkinson’s. For up-to-date information on the study, visit https://www.parkinson.org/pdgeneration.

## Author Roles

(1) Research Project: A. Conception, B. Organization, C. Execution; (2) Statistical Analysis: A. Design, B. Execution, C. Review and Critique; (3) Manuscript Preparation: A. Writing of the First Draft, B. Review and Critique.

L.V.C.: 1A, 1B, 1C, 2A, 2B, 3A

S.C.P.: 2A, 2B, 3B

E.N.S.: 2B, 3B

L.L.: 2B, 3B

E.Y.: 2B, 3B

F.A.: 2B, 3B

J.A.: 2B, 3B

M.T.: 2B, 3B

J.A.R.: 2B, 3B

C.W.: 2B, 3B

O.M.: 2B, 3B

Y.D.: 2B, 3B

N.D.: 2B, 3B

A.T.: 2B, 3B

A.E.: 2B, 3B

S.P.: 2B, 3B

I.M.: 2B, 3B

L.G.: 2B, 3B

S.H.B.: 2B, 3B

O.G.: 2B, 3B

M.R.: 2B, 3B

P.B.: 2B, 3B

C.B.: 2B, 3B

A.A.D.: 2B, 3B

J.C.B.: 2B, 3B

K.S.: 2B, 2C, 3B

C.K.: 2B, 3B

R.N.A.: 2B, 3B

Z.G.O.: 1A, 1B, 2A, 3B

## Financial Disclosures of All Authors of the Preceding 12 Months

L.V.C., S.C.P., E.N.S., L.L., E.Y., F.A., J.A., M.T., J.A.R., C.W., O.M., Y.D., N.D., A.T., A.E., S.P., I.M., L.G., S.H.B., O.G., M.R., P.B., C.B., A.A.D., J.C.B., K.S., C.K., and R.N.A. have nothing to report.

## Data Availability Statement

All data generated in this study, including the variants analyzed, are available within the main manuscript. The analysis code is publicly accessible at https://github.com/gan-orlab/GBA-p.E388K-Meta-analysis. Portions of the McGill cohorts are available through the Canadian Open Parkinson Network (C-OPN), with access to genetic and related data granted upon request and approval by the C-OPN data access committee (https://copn-rpco.ca/submit-a-request/). AMP-PD data were accessed and analyzed via the Terra platform (https://amp-pd.org/), and UK Biobank data were obtained as whole-genome sequencing datasets through the UK Biobank Research Analysis Platform (https://www.ukbiobank.ac.uk/).

## Notes

**Relevant conflicts of interest/financial disclosures:** Z.G.-O. has received consulting fees from Lysosomal Therapeutics Inc. (LTI), Idorsia, Prevail Therapeutics, Inceptions Sciences (now Ventus), Neuron23, Handl Therapeutics, UCB, Ono Therapeutics, Denali, Bial Biotech, Capsida, Takeda, Simcere, EG427, Jazz Pharmaceuticals, Guidepoint, Lighthouse, and Deerfield. CK has received consulting fees from Centogene and Biogen, speakers’ honoraria from Bial, and royalties from Oxford University Press and Springer Nature.

**Funding agencies:** This study was financially supported by grants from the Michael J. Fox Foundation, the Canadian Consortium on Neurodegeneration in Aging (CCNA), the Canada First Research Excellence Fund (CFREF), awarded to McGill University for the Healthy Brains for Healthy Lives initiative (HBHL), and Parkinson Canada. The Columbia University cohort is supported by the Parkinson’s Foundation, the National Institutes of Health (K02NS080915 and UL1TR000040) and the Brookdale Foundation. We also recognize the contributions of the G-Can (GBA1-Canada) Initiative, an open-science partnership focused on research into GBA1-related neurodegeneration. G-Can receives support from the Galen and Hilary Weston Foundation, the Silverstein Foundation, and J. Sebastian van Berkom and Ghislaine Saucier

### Competing Interest Statement

Z. G.-O. received consultancy fees from Lysosomal Therapeutics Inc. (LTI), Idorsia, Prevail Therapeutics, Inceptions Sciences (now Ventus), Neuron23, Handl Therapeutics, UCB, Capsida, Vanqua Bio, Congruence Therapeutics, Ono Therapeutics, Denali, Bial Biotech, Bial, EG427, Takeda, Jazz Pharmaceuticals, Simcere, Guidepoint, Lighthouse, and Deerfield. K.S. received consultancy fees from Acurex

### Funding Statement

This study was financially supported by grants from the Michael J. Fox Foundation, the Canadian Consortium on Neurodegeneration in Aging (CCNA), the Canada First Research Excellence Fund (CFREF), awarded to McGill University for the Healthy Brains for Healthy Lives initiative (HBHL), and Parkinson Canada. The Columbia University cohort is supported by the Parkinsons Foundation, the National Institutes of Health (K02NS080915 and UL1TR000040) and the Brookdale Foundation. We also recognize the contributions of the G Can (GBA1 Canada) Initiative, an open science partnership focused on research into GBA1 related neurodegeneration. G Can receives support from the Galen and Hilary Weston Foundation, the Silverstein Foundation, and J. Sebastian van Berkom and Ghislaine Saucier

### Author Declarations

All participating institutions obtained approval for study protocols from their respective institutional review boards, and written informed consent was secured from all individuals before enrollment. Analyses were conducted under approval from the McGill Institutional Review Board (IRB protocol A11-M60-21A).

### Summary of Updates

Change in surname of one of the corresponding authors

