## Supplemental files for "The *GBA1* p.E427K (p.E388K) Variant is a Risk Factor for Synucleinopathies: A Meta-Analysis"

Parkinson's Foundation

PD GENEration

POWERED BY THE PARKINSON'S FOUNDATION

### Supplementary Material: PD GENEration Affiliations for Banner Authorship

This supplemental document provides a list of affiliated members of the PD GENEration study under banner authorship for the associated manuscript as per the PD GENEration publication policy (*Rev 1/26*). Affiliations reflect institutional membership at the time of submission.

| PDGENE Affiliation | Name | Affiliation(s) |
|---|---|---|
| Local Site Investigator | Pinky Agarwal, MD | Evergreen Health, Seattle, WA |
| National Recruiting Site Investigator; Local Site Investigator | Julian Agin-Liebes, MD | Columbia University, New York, NY |
| Principal Investigator; Local Site Investigator; Steering Committee | Roy N. Alcalay, MD, MS | Tel Aviv Sourasky Medical Center, Tel Aviv, Israel<br>Columbia University, New York, NY |
| LATAM Site Investigator | Sarael Alcauter, PhD | Universidad Autónoma de México, Santiago de Querétaro, QRO, Mexico |
| Local Site Investigator | Jason Aldred, MD | Inland Northwest Research, Spokane, WA |
| Core Team | Ignacio Azcarate | Parkinson's Foundation, New York, NY |
| Local Site Investigator | Matthew Barrett, MD | Virginia Commonwealth University, Richmond, VA |
| Study Investigator; Steering Committee; Core Team | James C. Beck, PhD | Parkinson's Foundation, New York, NY |
| Local Site Investigator | Karen Blindauer, MD | Medical College of Wisconsin, Milwaukee, WI |
| Core Team | Nicola Bothwick | Parkinson's Foundation, New York, NY |
| Local Site Investigator | Chantale Branson, MD | Morehouse College, Atlanta, GA |
| Local Site Investigator | Susan Bressman, MD | Mount Sinai Beth Israel Medical Center, New York, NY |
| National Recruiting Site Investigator; Local Site Investigator | Jeff Bronstein, MD, PhD | University of California Los Angeles (UCLA), Los Angeles, CA |
| Local Site Investigator | Michiko Bruno, MD | Queen's Health System, Honolulu, HI |
| Core Team | Lark Caboy | Parkinson's Foundation, New York, NY |
| Genetics Counseling Core | Valentina Caceres, MS, CGC | Indiana University, Indianapolis, IN |

| PDGENE Affiliation | Name | Affiliation(s) |
|---|---|---|
| LATAM Site Investigator | Pedro Chaná, MD | Corporación Centro de Trastornos del Movimiento, Santiago, Chile |
| Local Site Investigator | Kelvin Chou, MD | University of Michigan, Ann Arbor, MI |
| Genetics Counseling Core | Lola Cook, MS, CGC3 | Indiana University, Indianapolis, IN |
| LATAM Site Investigator | Mario Cornejo Olivas, MD | Instituto Nacional de Ciencias Neurológicas, Lima, Perú |
| LATAM Site Investigator | Rossy Cruz Vicioso, MD | Unión Médica, Clínica Universitaria, Santiago de los Caballeros, Dominican Republic |
| Local Site Investigator | Nabila Dahodwala, MD | University of Pennsylvania, Philadelphia, PA |
| Local Site Investigator | Tom Davis, MD | Vanderbilt University, Nashville, TN |
| Core Team | Rebeca De Leon | Parkinson's Foundation, New York, NY |
| Local Site Investigator | Joy Antonelle de Marcaida, MD | Hartford HealthCare, Hartford, CT |
| Local Site Investigator | Marissa Dean, MD | University of Alabama Birmingham, Birmingham, AL |
| Local Site Investigator | Amanda Deligtisch, MD | University of New Mexico, Albuquerque, NM |
| Local Site Investigator | Rohit Dhall, MD | University of Arkansas, Fayetteville, AR |
| Core Team | Allison A. Dilliott, PhD | Parkinson's Foundation, New York, NY |
| Core Team | Megan Dini, MA | Parkinson's Foundation, New York, NY |
| Local Site Investigator | Elizabeth Disbrow, PhD | Louisiana State University, Baton Rouge, LA |
| Core Team | Kirby Doshier, PhD | Parkinson's Foundation, New York, NY |
| LATAM Site Investigator | Ingrid Estmann, MD | Universidad Autónoma de Nuevo León, San Nicolás de los Garza, Mexico |
| LATAM Site Investigator | Elias Fernandez, MD | Universidad de Concepción, Concepción, Chile |
| Local Site Investigator | Hubert Fernandez, MD | Cleveland Clinic, Cleveland, OH |
| Local Site Investigator | Jeanne Feuerstein, MD | University of Colorado, Boulder, CO |
| Local Site Investigator | Nicholas Fleming, MD | Atrium Health, Charlotte, NC |
| Genetics Counseling Core | Katherine Fiallos, CGC3 | Indiana University, Indianapolis, IN |
| Core Team | Megan Finke | Parkinson's Foundation, New York, NY |
| Steering Committee; National Recruiting Site Investigator; Genetics Counseling Core | Tatiana Foroud, PhD, | Indiana University, Indianapolis, IN |

Parkinson's Foundation

PD GENEration

POWERED BY THE PARKINSON'S FOUNDATION

| PDGENE Affiliation | Name | Affiliation(s) |
|---|---|---|
| Genetics Testing Core | Harry Gao, PhD | Fulgent Genetics, Temple City, CA |
| Steering Committee; PDGENE Team Lead | Kamalini Ghosh Galvelis, MS | Parkinson's Foundation, New York, NY |
| Local Site Investigator | Ro'ee Gilron, PhD | Rune Labs, San Francisco, CA |
| Local Site Investigator | Steven Gunzler, MD | Case Western, Cleveland, OH |
| Past Steering Committee | Anne Hall, MD | Parkinson's Foundation, New York, NY |
| Local Site Investigator | Deborah Hall, MD | Rush University, Chicago, IL |
| Local Site Investigator | Ihtsham Haq, MD | University of Miami, Miami, FL |
| Local Site Investigator | Sharon Hassin, MD | Chaim Sheba Medical Center, Ramat Gan, Israel |
| Genetics Counseling Core | Laura Heathers | Indiana University, Indianapolis, IN |
| Local Site Investigator | Emily Hill, MD | University of Cincinnati, Cincinnati, OH |
| Genetics Counseling Core | Priscila Hodges, MS, CGC3 | Indiana University, Indianapolis, IN |
| Local Site Investigator | Anna Hohler, MD | BMC Community Hospital, Boston, MA |
| Local Site Investigator | Stuart Isaacson, MD | Parkinson's Disease & Movement Disorders Center of Boca Raton, Boca Raton, FL |
| LATAM Site Investigator | Marcelo Kauffman, MD, PhD | Hospital Ramos Mejia, Buenos Aires, Argentina |
| Local Site Investigator | Tarannum Khan, MD | Cleveland Clinic Weston, Weston, FL |
| Local Site Investigator | Yasaman Kianirad, MD | University of Illinois-Chicago, Chicago, IL |
| Local Site Investigator | Annie Killoran, MD | University of Iowa, Iowa City, IA |
| Local Site Investigator | Sushma Kola, MD | Allegheny Health Network, Pittsburg, PA |
| Site and Clinical Data Management Core; Steering Committee | Sarah Lawrence, MS | Navitas Clinical Research, Rockville, MD |
| Site and Clinical Data Management Core | Susan Li, MD, PhD | Navitas Clinical Research, Rockville, MD |
| Local Site Investigator | Tsao-Wei Liang, MD | Thomas Jefferson University, Philadelphia, PA |
| Local Site Investigator | Irene Litvan, MD | University of California San Diego, La Jolla, CA |
| Site and Clinical Data Management Core | Yun Lu, PhD | Navitas Clinical Research, Rockville, MD |

| PDGENE Affiliation | Name | Affiliation(s) |
|---|---|---|
| Local Site Investigator | Irene Andonia Malaty, MD | University of Florida, Gainesville, FL |
| Past Steering Committee | Karen S. Marder, MD, MPH | Columbia University, New York, NY |
| Local Site Investigator | Zoltan Mari, MD | Cleveland Clinic Las Vegas, Las Vegas, NV |
| Local Site Investigator | Connie Marras, MD, PhD | Toronto Western Hospital, Toronto, ON |
| Steering Committee | Ignacio Mata, PhD | Cleveland Clinic, Cleveland, OH |
| Local Site Investigator | Kathleen McKee, MD | Intermountain Healthcare, Salt Lake City, UT |
| Steering Committee; National Recruiting Site Investigator; Local Site Investigator | Niccolo Mencacci, MD, PhD | Northwestern University, Chicago, IL |
| Genetics Testing Core | Yan Meng, PhD | Fulgent Genetics, Temple City, CA |
| Genetics Counseling Core | Amanda Miller, MS, CGC3 | Indiana University, Indianapolis, IN |
| Local Site Investigator | Kelly Mills, MD | Johns Hopkins University, Baltimore, MD |
| Local Site Investigator | Janis Miyasaki, MD | University of Alberta, Edmonton, AB |
| Steering Committee; Local Site Investigator | Martha Nance, MD | Park Nicollet Struthers Parkinson's Center, Minneapolis, MN |
| Genetics Counseling Core | Alia Neibaur | Indiana University, Indianapolis, IN |
| Core Team | Melissa Nicewaner | Parkinson’s Foundation, New York, NY |
| National Recruiting Site Investigator; Local Site Investigator | Eleni Okeanis Vaou, MD | University of Texas San Antonio (UTSA), San Antonio, TX |
| LATAM Site Investigator | Jorge Luis Orozco Vélez, MD | Fundación Valle del Lili, Cali, Colombia |
| Core Team | Sarah Osborne | Parkinson’s Foundation, New York, NY |
| Local Site Investigator | Jill Ostrem, MD | University of California San Francisco, San Francisco, CA |
| Local Site Investigator | Rajesh Pahwa, MD | University of Kansas, Lawrence, KS |
| Local Site Investigator | Gian Pal, MD | Rutgers University, New Brunswick, NJ |
| LATAM Site Investigator | Dr. Floria carla Pancetti Vaccari | Universidad Católica del Norte, Coquimbo, Chile |
| National Recruiting Site Investigator; Local Site Investigator | Ariane Park, MD | Ohio State University Medical Center, Columbus, OH |

| PDGENE Affiliation | Name | Affiliation(s) |
|---|---|---|
| LATAM Site Investigator | Susana Lissette Peña Martinez, MD | Universidad Dr. Andrés Bello, San Salvador, El Salvador |
| Steering Committee | John Poma | Parkinson’s Foundation, New York, NY |
| Local Site Investigator | Joseph Quinn, MD | Oregon Health & Science University, Portland, OR |
| Site and Clinical Data Management Core | Uma Ragunathan, MS | Navitas Clinical Research, Rockville, MD |
| Local Site Investigator | Gonzalo Revuelta, DO | Medical University of South Carolina, Charleston, SC |
| Local Site Investigator | Giulietta Riboldi, MD | New York University, New York, NY |
| Local Site Investigator | Bernardo Rodrigues, MD PhD | University of Connecticut, Mansfield, CT |
| LATAM Site Investigator | Mayela Rodríguez Violante, MD | Instituto Nacional de Neurología y Neurocirugía, Ciudad de México, México |
| Core Team | Joshua Ruffner | Parkinson’s Foundation, New York, NY |
| Genetics Counseling Core | Malia Rumbaugh | Indiana University, Indianapolis, IN |
| LATAM Site Investigator | Paula Saffie Awad, MD, PhD | Clínica Santa María, Santiago, Chile |
| Local Site Investigator | Julie Schwartzbard, MD | Aventura Neurologists, Aventura, FL |
| Steering Committee | Michael Schwarzschild, MD, PhD | Massachusetts General Hospital, Boston, MA |
| Steering Committee | Ruth Schneider, MD | University of Rochester, Rochester, NY |
| Local Site Investigator | Holly Shill, MD | Barrow Neurological Institute, Phoenix, AZ |
| Local Site Investigator | Lisa Shulman, MD | University of Maryland, College Park, MD |
| Local Site Investigator | David Simon, MD, PhD | Beth Israel Deaconess Medical Center, Boston, MA |
| Past Steering Committee | Tanya Simuni, MD | Northwestern University, Chicago, IL |
| Local Site Investigator | Claudia Testa, MD, PhD | University of North Carolina - Chapel Hill, Chapell Hill, NC |
| Core Team | Max Thom | Parkinson’s Foundation, New York, NY |
| Genetics Counseling Core | Michelle Totten | Indiana University, Indianapolis, IN |
| Local Site Investigator | Blanca Valdovinos, MD | University of Rochester, Rochester, NY |
| Local Site Investigator | Nora Vanegas, MD | Baylor College of Medicine, Houston, TX |

Parkinson's Foundation

PD GENEration

POWERED BY THE PARKINSON'S FOUNDATION

| PDGENE Affiliation | Name | Affiliation(s) |
|---|---|---|
| Genetics Counseling Core | Jennifer Verbrugge, MS, CGC3 | Indiana University, Indianapolis, IN |
| Past Steering Committee; Local Site Investigator | Anne-Marie Wills, MD | Massachusetts General Hospital, Boston, MA |
| Local Site Investigator | Rebecca Williamson, MD, PhD | University of Pennsylvania, Philadelphia, PA |
| Local Site Investigator | Katherine Wong, MD | University of Southern California, Los Angeles, CA |
| Local Site Investigator | Tao Xie, MD, PhD | University of Chicago, Chigaco, IL |
| Local Site Investigator | Gilad Yahalom, MD | Shaare Zedek Medical Center, Jerusalem, Israel |
| Core Team | Addison Yake | Parkinson's Foundation, New York, NY |
| Local Site Investigator | Tritia Yamasaki, MD, PhD | University of Kentucky, Lexington, KY |
| Core Team | Anny Coral Zambrano | Parkinson's Foundation, New York, NY |
| Local Site Investigator | Hengameh Zahed, MD, PhD | Stanford University, Stanford, CA |
| Local Site Investigator | Elizabeth Zauber, MD | Indiana University, Indianapolis, IN |

*Alphabetical order by last name.*
